# Evaluation of diagnostic performance of the “STANDARD G6PD™” quantitative point-of-care test in neonates and infants

**DOI:** 10.64898/2026.03.26.26349364

**Authors:** Gornpan Gornsawun, Eh Moo, Klay Htoo, Supalak Chalermvisutkul, Marie Ellen Gilder, Phaw Koo Mu, Laypaw Archusuksan, Taco Jan Prins, Borimas Hanboonkunupakarn, Rose McGready, Francois Nosten, Germana Bancone

## Abstract

Severe neonatal hyperbilirubinaemia represents a considerable cause of mortality and long term-morbidity in neonates born in low resource settings. Early identification of risk factors, such as glucose-6-phosphate dehydrogenase (G6PD) status, has the potential to prevent severe hyperbilirubinaemia and improve the clinical outcomes.

The primary aim of the study was to assess equivalency of cord blood and neonatal capillary blood for diagnosis of G6PD deficiency using the quantitative point-of-care “STANDARD G6PD™” test (SD Biosensor, Korea). Additional secondary aims were to compare the “STANDARD G6PD™” with gold standard spectrophotometry and to analyse changes in G6PD activity in the first 4 months of life.

A total of 75 neonates born in Shoklo Malaria Research Unit (SMRU) clinics were selected based on their G6PD status assessed through routine cord blood screening using the “STANDARD G6PD™” test. Using activity thresholds established before in this setting, 25 G6PD deficient, 25 G6PD intermediate and 25 G6PD normal neonates were identified and re-tested on capillary blood collected within 24 hours of life and at day 7. They were also followed-up at 1 and 4 months of age to study haematologic and G6PD activity changes over time.

The results showed that the “STANDARD G6PD™” can be used reliably up to one week of life for testing neonates using the same thresholds established in cord blood. Performance of the point-of-care test as compared to the gold standard spectrophotometry remained excellent at all sampling time-points. Nevertheless, G6PD activity assessed longitudinally in the same participants decreased over time, both at 1 month of age and at 4 months of age, and interpretation of results in female infants with intermediate activity might require different thresholds.

The study demonstrated that the “STANDARD G6PD™” can effectively support clinical care in neonates and infants in populations with prevalent G6PD deficiency at the primary care level and especially in low-resource settings.

## Introduction

Bilirubin is a byproduct of haemoglobin catabolism. Levels of bilirubin are generally high in neonates (physiological hyperbilirubinaemia) due to increased destruction of fetal red blood cells and reduced capacity of immature enterohepatic systems to conjugate and excrete bilirubin. While physiological hyperbilirubinaemia is a normal finding in healthy neonates and generally does not require treatment, pathological neonatal hyperbilirubinaemia (NH) can lead to an accumulation of unconjugated bilirubin in the brain causing life-long neurological sequelae (kernicterus) and even death (Slusher, Zamora et al. 2017, Wouda, Thielemans et al. 2020). Glucose-6-phosphate dehydrogenase (G6PD) deficiency is one of the major risk factors for NH (Kaplan and Hammerman 2002). Phototherapy in equipped clinics is the mainstay of treatment but more severe NH might require exchange transfusion. Early diagnosis of abnormal G6PD phenotypes can help provide appropriate clinical care, both by avoiding treatment with possibly haemolytic drugs but also by extending observation and repeated bilirubin testing of neonates with this risk factor. Extended clinical care at birthing centers can dramatically improve outcomes in mobile and vulnerable populations with difficult access to care (Olusanya, Osibanjo et al. 2015). If not done at birth, diagnosis of G6PD status is often sought during the first week of life in neonates presenting with hyperbilirubinaemia and during the first month of life in neonates with prolonged jaundice.

Reference normal ranges of G6PD enzymatic activity have been established in SMRU in cord blood samples collected at birth (Bancone, Gilder et al. 2022) and have been used since to identify neonates most at risk. Nevertheless, around 15% of neonates in this setting are not tested at birth using cord blood because they are born outside the clinic, and these neonates are tested at a later moment. It is important to confirm whether data obtained in cord blood are directly comparable to capillary whole blood collected within the first 24 hours of life or later. Published data indicate that there is a gradual decrease in enzymatic activity over time during the first 12 months of life (Travis, Kumar et al. 1980, Tang, Liu et al. 1995, Yang, Tai et al. 2019, Bancone, Poe et al. 2022) but there are no data on the changes in the first week and first month of life when the test is most often performed and large haematologic changes are observed. From the clinical point of view, it is important to understand whether G6PD activity thresholds established in cord blood can be used for the first week and month of life to diagnose neonates with abnormal G6PD levels.

The neonatal population in SMRU clinics shows high rates of NH (24% overall, (Turner, Carrara et al. 2013)) which is multifactorial (Thielemans, Peerawaranun et al. 2021) and has strong genetic components (Bancone, Gornsawun et al. 2022). In particular, G6PD deficiency which is very common among the Karen and Burman population attending SMRU clinics (15-20% in males, (Bancone, Chu et al. 2014)), is associated with an increased risk to develop NH requiring phototherapy in neonates with deficient and intermediate G6PD levels (Bancone, Gornsawun et al. 2022). The “STANDARD G6PD ™” test (SD Biosensor, Korea; henceforth referred as Biosensor) is a quantitative enzymatic colorimetric assay that measures G6PD enzyme activity normalized by haemoglobin on 10 μl of capillary or venous blood. Results expressed in units per gram of haemoglobin (U/gHb) are provided within two minutes on a portable, handheld analyzer and are used to classify individuals as G6PD normal, intermediate, or deficient according to established thresholds. In a previous evaluation study, the test has shown to provide an accurate diagnosis of G6PD deficiency when used on cord blood at birth (Bancone, Gilder et al. 2022). In SMRU it is routinely used to improve the clinical management of neonates at risk of NH and establishing its accuracy in capillary blood taken in the first month of life would extend these benefits to babies born outside SMRU clinics.

This study investigated whether levels of G6PD in cord blood and capillary neonatal blood are equivalent by Biosensor. Changes in G6PD activity in the first week and months of life in the same neonates followed-up prospectively until the 4-months were also analysed.

## Materials and Methods

Pregnant women of Karen and Burman ethnicity attending Antenatal care (ANC) at SMRU Maw Ker Thai (MKT) clinic on the Thailand-Myanmar border were approached to participate in the study. In late pregnancy, eligibility (age 18 years or older with a singleton pregnancy) was assessed and written informed consent signed by the mother.

In SMRU clinics, cord blood samples are routinely collected in 2ml EDTA tubes (after birth of the neonate and before delivery of the placenta) and analysed for G6PD activity using the Biosensor. The Biosensor test is performed by locally trained laboratory technicians following manufacturer’s instructions. Two level quality control samples are analysed weekly. Neonates are classified based on thresholds established before: deficient (≤4.8 IU/gHb), intermediate (4.9-9.9IU/gHb, females only) and normal (≥10.0IU/gHb). Counselling and a multi-language red card indicating the baby G6PD status and substances to avoid are given to mothers of neonates with abnormal G6PD levels.

For the study, eligibility of neonates was assessed after results of G6PD were available from the site laboratory. Neonates were included if they were born at an estimated gestational age (EGA) by ultrasound ≥35 weeks with no severe maternal complications at birth and no severe neonatal illness, and had G6PD activity within the aforementioned thresholds.

The sampling plan is presented in Figure 1.

**Figure 1.**
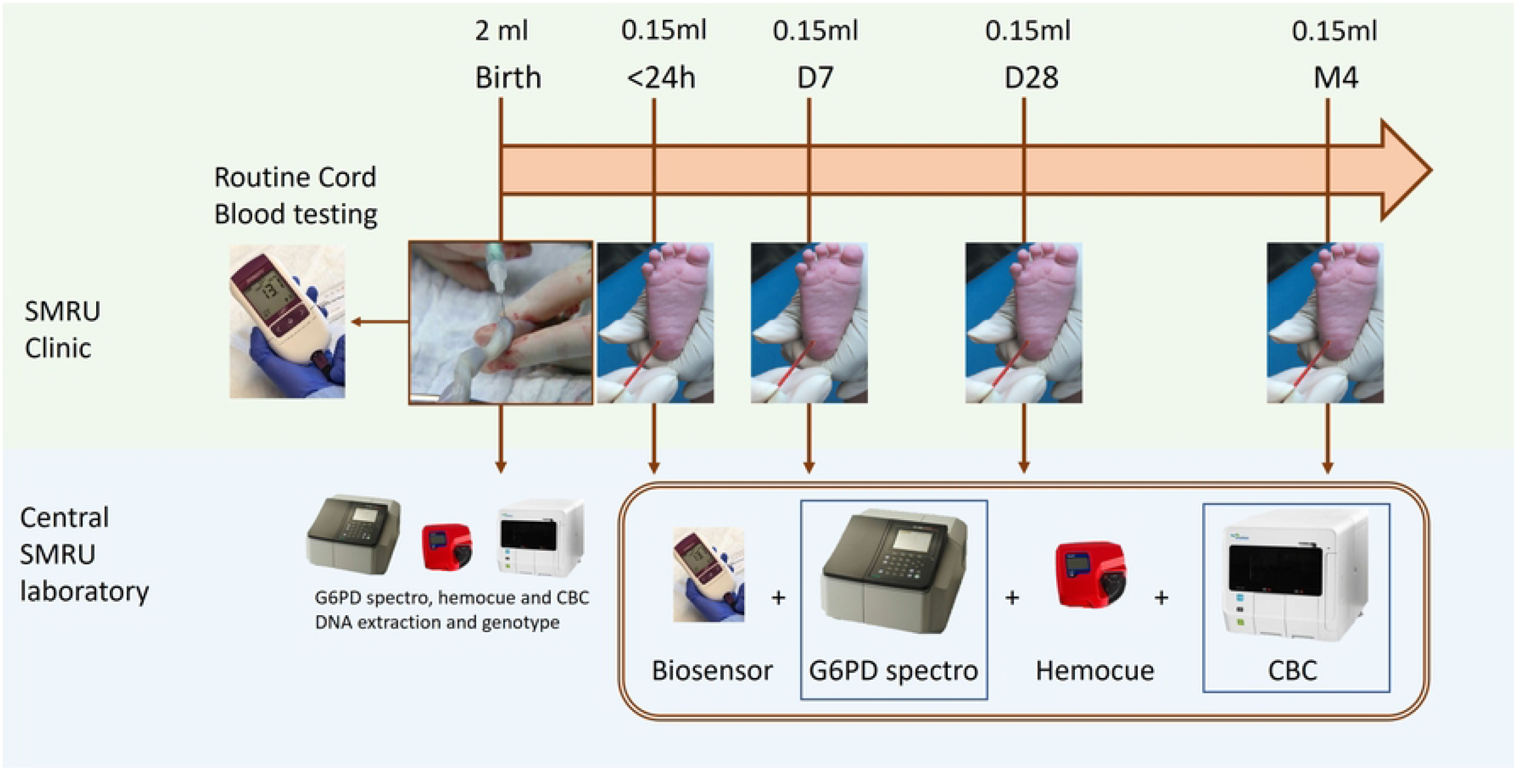
Study sampling plan.

Cord blood samples of participating neonates were transported on ice packs to the central haematology laboratory of SMRU in Mae Ramat within 24 hours and further analysed by reference spectrophotometric assay (Pointe Scientific assay kit # G7583-180, lysis buffer # G7583-LysSB) on a SHIMADZU UV-1800 spectrophotometer (SHIMADZU, Japan) with temperature-controlled compartment. Samples were also analysed by complete blood count, including reticulocyte count, using a Sysmex XN-550 haematology analyzer (Sysmex Thailand Co., Ltd) and by Hemocue 301+ (HemoCue AB, based in Ängelholm, Sweden). Laboratory operators were blind to Biosensor clinic results.

At follow-up, a heel-prick sampling of 150uL of capillary blood on EDTA was carried out within 24 hours from birth, at day 7, at day 28 and at 4 months of life and transported to central haematology laboratory to be analysed by Biosensor, spectrophotometric assay, complete blood count, and Hemocue 301+. At the haematology laboratory, operators of Biosensor testing were blind to spectrophotometric results.

Genotyping for G6PD mutations following SMRU SOPs and protocol from Kim and colleagues (Kim, Nguon et al. 2011) was performed from buffy coat obtained from cord blood. Genotype for Mahidol (487G>A), the most common G6PD mutation in this population, was performed first on all samples. Deficient and intermediate samples without Mahidol mutation, were then analysed for a panel of other common mutations (Bancone, Chu et al. 2014): Viangchan (871G>A), Chinese-4 (392G>T), Canton (1376G>T), Kaiping (1388G>A), Union (1360C>T), Mediterranean (563C>T). Full gene sequence was performed if none of these mutations were found.

Routine clinical care was provided to all neonates included in the study. After birth, a routine pre-discharge total serum bilirubin test was carried out around 12-48 hours after birth, depending on clinical and risk factors. When neonates developed NH during the first week of life, they were admitted to the Special Care Baby Unit (SCBU) of SMRU and treated with phototherapy as per SMRU clinical guidelines. The gestational age at birth, assessed by early pregnancy ultrasound, were categorised as ≤38 and >38 weeks according to epidemiological studies conducted previously in the same population (Thielemans, Peerawaranun et al. 2021).

### Sample size and statistical analyses

The sample size to assess equivalence was calculated considering a Pearson’s correlation (r) between G6PD in cord blood and capillary blood of 0.95. Therefore, assuming a 95% confidence interval and acceptable half-width interval 0.05, the minimum sample size needed was 21; to account for 10% attrition, 25 neonates per class of phenotype (deficient, intermediate and normal) were included for a total of 75 samples.

For the analysis of diagnostic performance, G6PD status was defined using Biosensor and spectrophotometry thresholds established before in cord blood (Bancone, Gilder et al. 2022). In order to understand whether universal thresholds could be used to define G6PD status at birth on cord blood and at follow-up visits, participants were classified as deficient, intermediate and normal based on Biosensor’s thresholds used in routine clinical screening; the classification was then applied to activity detected in the first capillary sample, at day 7, at day 28 and at month 4 capillary samples. The same was done with data obtained using the gold standard assay, using spectrophotometry-defined thresholds. These analyses were carried out only on participants with samples collected at all time-points.

Mean and SD or median and min-max values were reported for continuous variables. Correlation between activity in cord blood and at follow-up was assessed using Pearson’s coefficient of correlation. Difference in G6PD activity and blood indices in paired samples were analysed by Wilcoxon Signed-Rank Test and paired T-test respectively.

Intraclass Correlation coefficient and Bland-Altman plot were used to inspect correspondence between G6PD activity detected by Biosensor compared with the spectrophotometry assay. Bland-Altman analysis was performed by calculating the difference between individual paired measurements from the Biosensor and the gold standard. These differences were then plotted against the average of the two methods for each sample. The 95% Limits of Agreement were calculated as mean difference ±1.96SD.

Analyses were performed in SPSSv29 (IBM Corp., Armonk, USA).

## Results

### Study population

Study enrolment started on July 1^st^ 2024 and was completed on April 20^th^ 2025. Complete follow-up until 4 months of life for the last participant was reached on August 20^th^ 2025. A total of 76 neonates were enrolled, with one excluded after the cord blood collection because the capillary sample collected within 24h of life could not be shipped to the central haematology laboratory for spectrophotometric analysis.

Neonates included in the study were 39 females and 36 males, with mean gestational age (min-max) of 39.1 (36.1-41.0) weeks. All the 75 participants were born by normal vaginal birth and had cord blood sampled, and had capillary blood collected within 24h after birth, at day 7, and at day 28. A capillary sample was taken at 4 months age for 95% (71/75) but one sample was clotted. Remaining participants were lost to follow-up.

Using biosensor to test cord blood samples, and applying activity thresholds used at the clinic, 26 neonates (3 females, 23 males) were deficient (activity ≤4.8U/gHb), 26 were female intermediate (activity between 4.9 and 9.9U/gHb) and 23 (10 females, 13 males) had normal G6PD status (activity ≥10.0U/gHb) at enrolment. Out of the 26 deficient samples, one sample (GCN-048) was later confirmed to be G6PD normal by spectrophotometric analysis on the same blood, on all subsequent blood samples (by both Biosensor and spectrophotometry) and by genotyping. Original logbook records and results recorded in the device were checked but no obvious explanation could be found for the incorrect result, suggesting a procedural error during testing or sample mis-identification. For this participant, results from cord blood sample only were excluded from analyses.

Using spectrophotometry to test cord blood samples and previously defined G6PD activity thresholds (100% normal activity=13.3U/gHb), 25 neonates were identified as deficient (≤4.0IU/gHb), 20 as intermediate (4.1-9.3IU/gHb) and 30 as G6PD normal (≥9.4IU/gHb) at enrolment.

The first capillary sample was collected at an average age (SD) of 20.4 (3.20) hours (Figure S1). The second capillary sample was collected at a median age (min-max) of 7 (6-9) days of life. The third and fourth capillary samples were collected at a median age (min-max) of 28 (26-59) days of life and 122 (112-159) days of life.

Overall results of G6PD activity according to phenotypic group using Biosensor and spectrophotometry are presented in Table 1 and 2. Among all the 295 follow-up samples analysed by Biosensor, one sample collected within 24 hours of life did not provide a usable result by Biosensor (G6PD=NA with Hb=HI) and Hemocue; complete blood count in the sample showed Hb= 24.1 g/dL, the highest haemoglobin result recorded in the study.

**Table 1.**
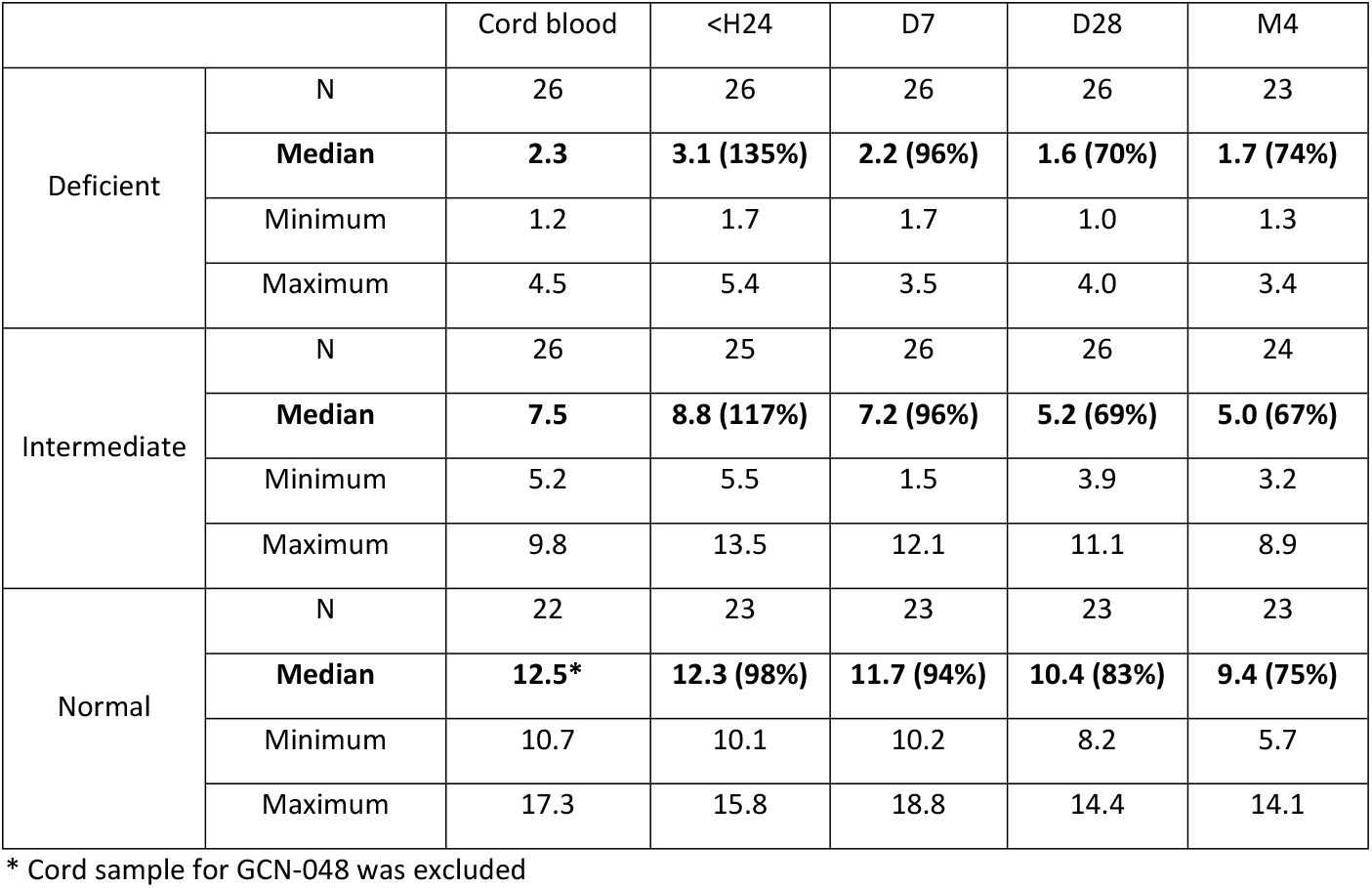
Median (minimum-max) G6PD activity by sampling day using the Biosensor (U/gHb) according to G6PD phenotype established at birth. Percentage of activity in follow-up samples as compared to cord blood is reported in brackets.

|  |  | Cord blood | <H24 | D7 | D28 | M4 |
| --- | --- | --- | --- | --- | --- | --- |
| Deficient | N | 26 | 26 | 26 | 26 | 23 |
|  | Median | 2.3 | 3.1 (135%) | 2.2 (96%) | 1.6 (70%) | 1.7 (74%) |
|  | Minimum | 1.2 | 1.7 | 1.7 | 1.0 | 1.3 |
|  | Maximum | 4.5 | 5.4 | 3.5 | 4.0 | 3.4 |
| Intermediate | N | 26 | 25 | 26 | 26 | 24 |
|  | Median | 7.5 | 8.8 (117%) | 7.2 (96%) | 5.2 (69%) | 5.0 (67%) |
|  | Minimum | 5.2 | 5.5 | 1.5 | 3.9 | 3.2 |
|  | Maximum | 9.8 | 13.5 | 12.1 | 11.1 | 8.9 |
| Normal | N | 22 | 23 | 23 | 23 | 23 |
|  | Median | 12.5* | 12.3 (98%) | 11.7 (94%) | 10.4 (83%) | 9.4 (75%) |
|  | Minimum | 10.7 | 10.1 | 10.2 | 8.2 | 5.7 |
|  | Maximum | 17.3 | 15.8 | 18.8 | 14.4 | 14.1 |

\* Cord sample for GCN-048 was excluded

**Table 2.**
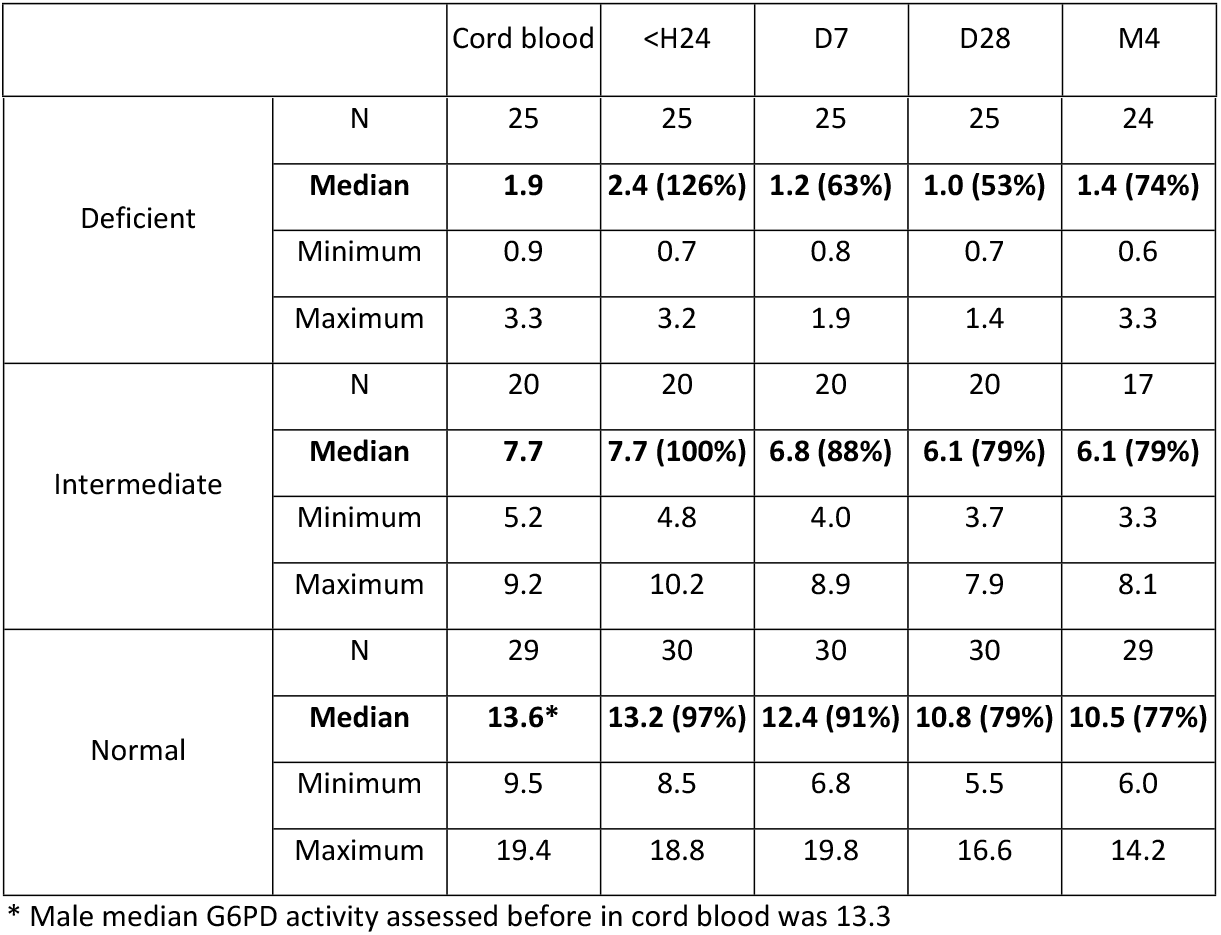
Median (min-max) G6PD activity by sampling day using the Spectrophotometer (IU/gHb) according to spectrophotometry-defined G6PD phenotype established at birth.

|  |  |  |  |  |  |  |
| --- | --- | --- | --- | --- | --- | --- |
| Deficient | N | 25 | 25 | 25 | 25 | 24 |
|  | Median | 1.9 | 2.4 (126%) | 1.2 (63%) | 1.0 (53%) | 1.4 (74%) |
|  | Minimum | 0.9 | 0.7 | 0.8 | 0.7 | 0.6 |
|  | Maximum | 3.3 | 3.2 | 1.9 | 1.4 | 3.3 |
| Intermediate | N | 20 | 20 | 20 | 20 | 17 |
|  | Median | 7.7 | 7.7 (100%) | 6.8 (88%) | 6.1 (79%) | 6.1 (79%) |
|  | Minimum | 5.2 | 4.8 | 4.0 | 3.7 | 3.3 |
|  | Maximum | 9.2 | 10.2 | 8.9 | 7.9 | 8.1 |
| Normal | N | 29 | 30 | 30 | 30 | 29 |
|  | Median | 13.6* | 13.2 (97%) | 12.4 (91%) | 10.8 (79%) | 10.5 (77%) |
|  | Minimum | 9.5 | 8.5 | 6.8 | 5.5 | 6.0 |
|  | Maximum | 19.4 | 18.8 | 19.8 | 16.6 | 14.2 |

\* Male median G6PD activity assessed before in cord blood was 13.3

Overall results of hemoglobin level by day of sampling by Biosensor, CBC and Hemocue are presented in Table S1.

G6PD Mahidol was the prominent mutation found with 20 heterozygous females, 21 hemizygous males and 1 homozygous female. Fourteen females and 13 males were wild type and other mutations were found only in single subjects: Viangchan, Coimbra, Acores and a new mutation 50G>A on exon 2 (causing an amino acid change from Arginine to Glutamine in position 17) were found in heterozygous females, while Kaiping and Orissa were found in hemizygous males. Table 3 reports the genotype distribution by G6PD phenotype as assessed by field Biosensor testing and laboratory-based spectrophotometry.

**Table 3.** Genotype according to G6PD status in cord blood (by Biosensor and Spectrophotometry)

| G6PD Status (CB) | G6PD genotype |  |  |
| --- | --- | --- | --- |
|  | Hemi/homozygote | Heterozygote | WT |
| Biosensor |  |  |  |
| Deficient | 24 | 2 | 0 |
| Intermediate | 0 | 22 | 4 |
| Normal | 0 | 0 | 23 |
| Spectrophotometry |  |  |  |
| Deficient | 24 | 1 | 0 |
| Intermediate | 0 | 20 | 0 |
| Normal | 0 | 3 | 26 |

### G6PD activity by Biosensor: comparison between cord blood and follow-up capillary samples

Pearson’s correlation coefficient between G6PD in cord blood and in capillary sample collected within 24h from birth was 0.937 (p<0.01) when analysed by Biosensor (Figure S2) and 0.980 (p<0.01) when analysed by spectrophotometer. G6PD activity in paired samples was significantly higher at 24 hours compared with cord blood in capillary blood of deficient participants when analysed by Biosensor or spectrophotometry and in intermediate participants analysed by Biosensor only. Activity was not different in G6PD normal participants analysed by either method (Table 1 and 2).

Pearson’s correlation coefficient between G6PD in cord blood and day 7 was 0.945 (p<0.01) when analysed by Biosensor (Figure S3) and was 0.968 (p<0.001) when analysed by spectrophotometer. Overall G6PD activity in paired samples was significantly lower in capillary blood from day 7 compared with cord blood when analysed by Biosensor (P=0.03) and by spectrophotometry (P<0.01). Since activity was expected to decrease after 1 month of life (Thielemans, Gornsawun et al. 2018) and after, no correlation analysis was performed for samples collected at day 28 and 4 months.

When comparing status classification, all G6PD deficient participants but one (a deficient heterozygote) would have been correctly identified as deficient by 24-hours and day-7 capillary blood using cord blood thresholds by Biosensor (Figure 2A) and spectrophotometry. All G6PD mutated hemizygotes and homozygotes would have been correctly identified as deficient by 24-hours and day-7 capillary blood using cord blood thresholds by Biosensor (Figure 2B). At 24 hours, there were 6 participants with intermediate activity who would be classified differently if analysis was carried out in capillary blood rather than cord blood (highlighted in red in Table 4, left pane) using Biosensor. At day 7, there were 5 G6PD intermediate participants who would have been classified differently using capillary blood (highlighted in red in Table 4, right pane) when analysed by Biosensor. When using spectrophotometry, two participants (1 intermediate and 1 normal) would have been classified differently using capillary blood within 24 hours as compared to cord blood (highlighted in red in Table 5, left pane) and 2 G6PD normal who would have been categorized differently using capillary blood at day 7 (Table 5, right pane).

**Figure 2.**
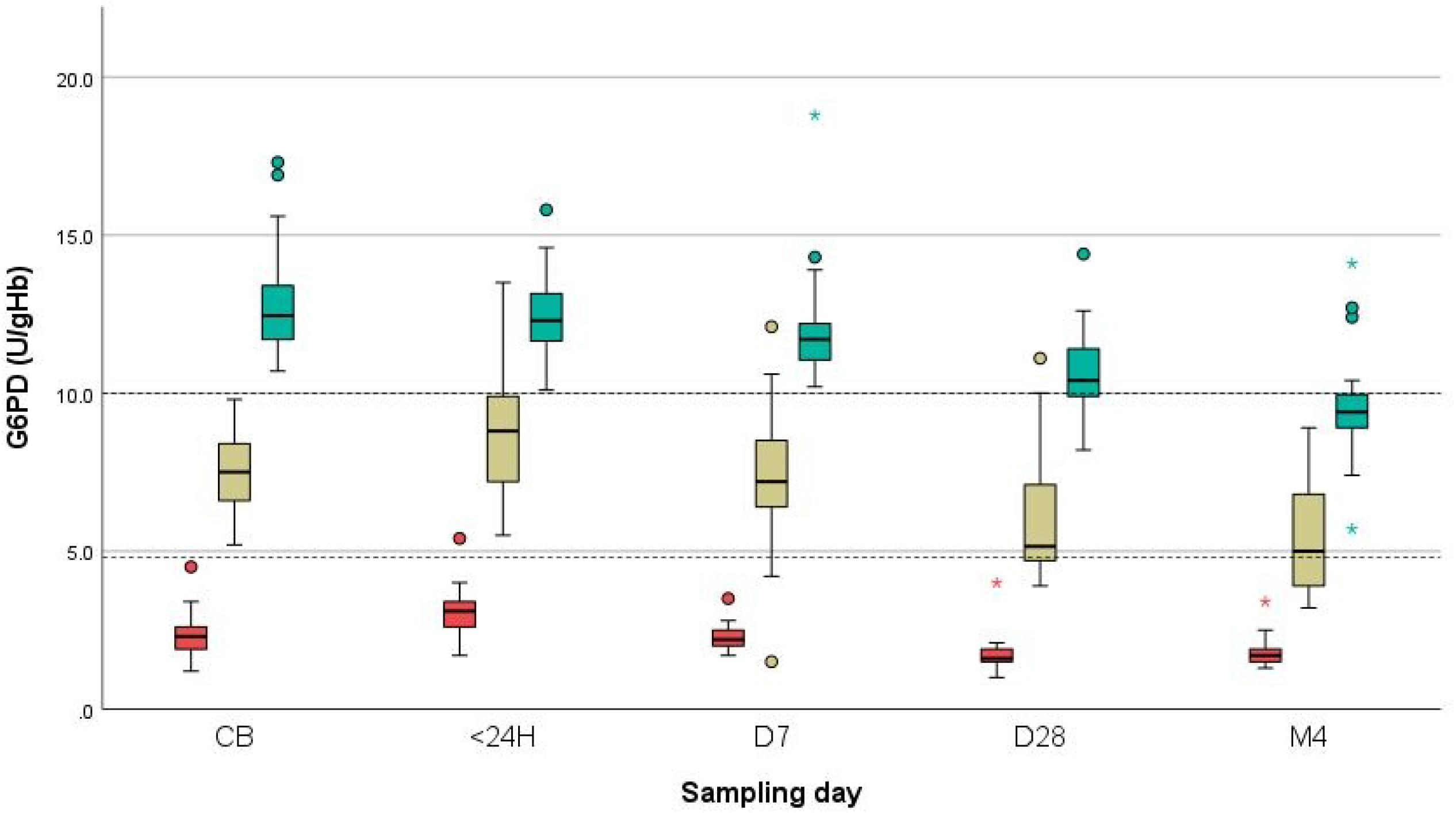
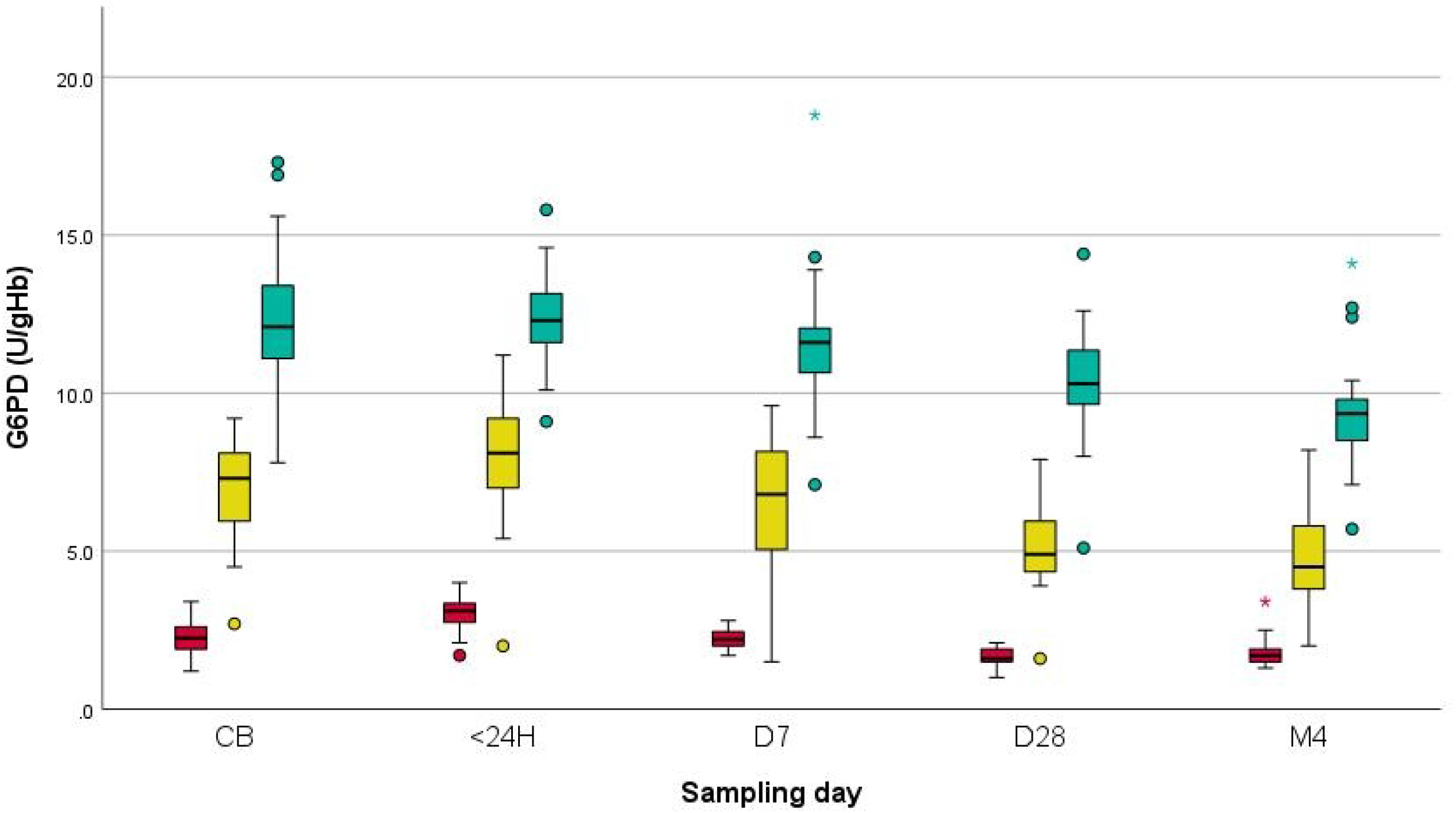
**(A) G6PD activity changes over time by Biosensor according to G6PD status established on cord blood.** Samples were classified as G6PD deficient (red bars), intermediate (yellow bars) and normal (green bars). Dotted lines indicate activity thresholds. **(B) G6PD activity changes over time by Biosensor according to genotype.** Hemizygote or homozygote mutated genotype (red bars), heterozygote (yellow) and wild type (green). CB: cord blood; <24H: sample collected within 24hours of life; D7: day 7; D28: day 28; M4: month 4.

**Table 4.** Comparison of G6PD status by Biosensor (using Biosensor CB thresholds)

|  |  | G6PD Status (<24H) |  |  | G6PD Status (D7) |  |  |
| --- | --- | --- | --- | --- | --- | --- | --- |
| G6PD status |  |  |  |  |  |  |  |
| Cord blood |  | Deficient | Intermediate | Normal | Deficient | Intermediate | Normal |
| Deficient* | 23 | 23 | 0 | 0 | 23 | 0 | 0 |
| Intermediate | 23 | 0 | 17 | 6 | 3 | 18 | 2 |
| Normal | 22 | 0 | 0 | 22 | 0 | 0 | 22 |

\* One G6PD heterozygote female with deficient phenotype not included here because she missed M4 follow-up had G6PD activity at 24H of 5.4U/gHb and would have been classified as intermediate. All subsequent G6PD tests were <4.8U/gHb threshold.

**Table 5.** Comparison of G6PD status by spectrophotometry (using spectrophotometric CB thresholds)

|  |  | G6PD Status (<24H) |  |  | G6PD Status (D7) |  |  |
| --- | --- | --- | --- | --- | --- | --- | --- |
| G6PD status |  |  |  |  |  |  |  |
| Cord blood |  | Deficient | Intermediate | Normal | Deficient | Intermediate | Normal |
| Deficient | 23 | 23 | 0 | 0 | 23 | 0 | 0 |
| Intermediate | 17 | 0 | 16 | 1 | 0 | 17 | 0 |
| Normal | 28 | 0 | 1 | 27 | 0 | 2 | 26 |

### G6PD activity and haemoglobin by Biosensor compared to gold standard spectrophotometry and CBC

ICC for G6PD activity between methods overall and by sampling day was excellent (Table S2). Mean difference in G6PD activity by sampling day and overall was generally very small (Table 6) but limits of agreement were wide. Bland-Altman graphs for comparison of all data points and by day of sampling are presented in Figure 3 and Figures S8-S12. Differences between methods were smallest at low activity levels and wider at higher activity levels.

**Table 6.** Mean differences G6PD activity (Biosensor – spectrophotometry) by sampling day.

|  | CB | H24 | D7 | D28 | M4 | Overall |
| --- | --- | --- | --- | --- | --- | --- |
| N | 74 | 73 | 74 | 74 | 68 | 367 |
| Mean | -0.75 | -0.14 | -0.16 | -0.31 | -0.75 | -0.42 |
| SD | 1.74 | 1.50 | 1.47 | 1.04 | 1.28 | 1.44 |
| LoA | -4.16 to 2.66 | -3.08 to 2.79 | -3.05 to 2.73 | -2.34 to 1.72 | -3.26 to 1.76 | -3.25 to 2.41 |

**Figure 3.**
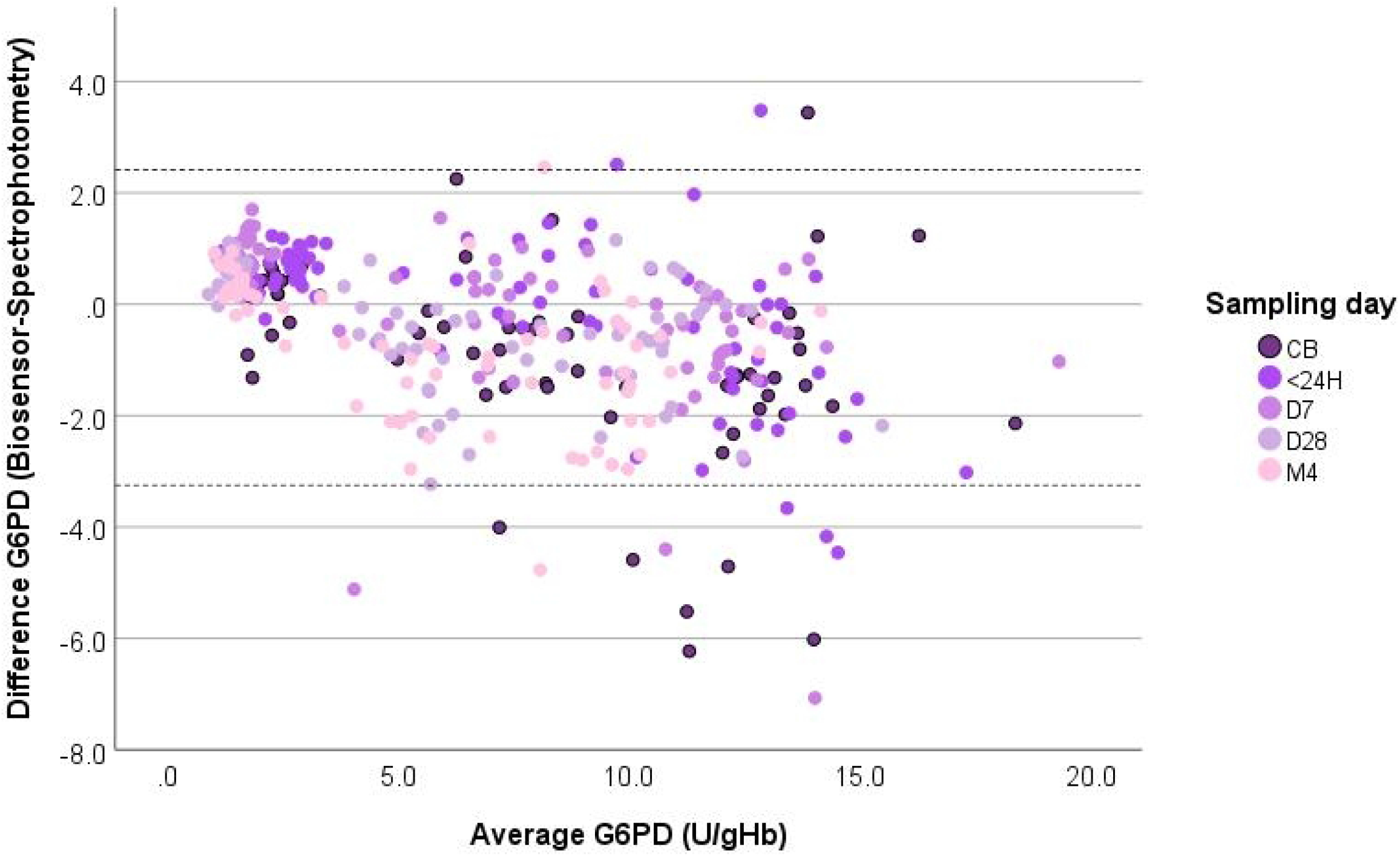
Bland-Altman plot for G6PD activity (IU/gHb) in Biosensor and spectrophotometry for all data points. Dotted lines represent limits of agreement.

For haemoglobin levels, overall ICC (95%CI) was 0.744 (0.17-9.07) between Biosensor and CBC and 0.86 (0.368-0.927) for Biosensor and Hemocue. Mean difference in haemoglobin between methods by sampling day and overall was generally acceptable (Table S3-S4) with very wide limits of agreement. Bland-Altman graphs for comparison of all data points is presented in Figure S13-S14 for CBC and Hemocue respectively.

### Other haematologic changes between cord blood and follow-up samples

Together with expected decrease in enzymatic activity, changes in G6PD activity detected during the first four months of life might be also influenced by other physiologic modifications in the number of white and red blood cells, reticulocytes and in the concentration of haemoglobin (Table S5 and Figure S4-S7). In particular, significant increase in WBC, RBC count and reticulocyte percentage at <24h of life in all G6PD phenotypic groups as compared to cord blood contributed to increased G6PD activity especially in G6PD abnormal participants.

### G6PD and neonatal hyperbilirubinaemia

Among neonates born at >38 weeks of gestational age, a higher proportion of G6PD deficient and intermediate neonates needed phototherapy (8/22 and 1/24 respectively) as compared to G6PD normal (0/21). When G6PD activity was analysed by genotype, 7/20 hemi and homozygotes and 2/22 heterozygotes needed phototherapy as compared to 0/24 wild types.

## Discussion

This study demonstrated that G6PD activity measured from cord blood and capillary blood collected up to 7 days of life can be used with the same thresholds to identify, reliably, G6PD deficient neonates in this setting. The majority but not all of G6PD female neonates with intermediate activity could be identified using the same thresholds. Longitudinal analyses further confirmed the expected physiologic decline of G6PD activity during the first months of life across phenotypic and genotypic groups. The study also confirmed the good performance of the Biosensor compared to the gold standard spectrophotometry in neonates and infants up to 4 months of life.

G6PD activity follows a clear bimodal distribution in males which allows a very good distinction between the normal and deficient phenotypes; the continuous distribution observed in females makes it more complicated to establish clear-cut thresholds and, consequently, performance of diagnostic tests (including the gold standard) at these thresholds are generally poorer in females. The current thresholds used in clinical practice are derived from physiologic and clinical observations with 30% activity as the “natural” threshold to define deficiency (corresponding to the flex in the male bimodal distribution) and the 70% activity as the conventional (but arbitrary to some degree) threshold to identify “normality” in females. The limitations of establishing thresholds for a continuous variable are clear but so is the need to have a practical indication for use in clinical routine practice, especially at the point-of-care. The “STANDARD G6PD” Biosensor test has been validated before in older children and adults in the context of malaria testing and specifically in *P. vivax* endemic regions to support use of 8-aminoquinolines for radical cure. Diagnostic performance of thresholds indicated by manufacturers (≤4.0U/gHb for deficiency and >6.0 for normality in females) have been evaluated in several studies with excellent results (Zobrist, Brito et al. 2021, Adissu, Brito et al. 2023, Martinez, Velez-Marin et al. 2024). It is acknowledged that both thresholds are conservative to maximise safety of 8-aminoquinolines administration.

Only 2 studies so far have analysed performance of the “STANDARD G6PD” test in neonates in cord blood or neonatal blood. The study from Manowong and colleagues (https://he01.tci-thaijo.org/index.php/CMMJ-MedCMJ/article/download/256783/173316/992256) assessed G6PD activity in venous blood from 76 G6PD normal male neonates (aged 1-7 days) establishing a male median of 12.7U/gHb with a proposed threshold to identify deficiency at 3.6 U/gHb; graphs of G6PD activity showed a clear bimodal distribution in males with a flex around 5U/gHb. A second study, performed previously in this setting, detected a male median by Biosensor of 14.4U/gHb and a threshold calculated to have the best sensitivity and specificity for genotypically hemi/homozygote participant at 4.8U/gHb. Table S6 shows results obtained before and data collected during this study; it includes a comparison with the manufacturer-defined thresholds established and validated in older children and adults in previous studies. Data obtained with the current study on a smaller sample size provided a median activity in G6PD wild type male and female participants of 12.1U/gHb in cord blood.

Follow-up samples showed a similar median G6PD activity up to day 7 (95% of cord) with activity declining to around 80% at 1 month as compared to cord blood and around 75% at month 4. Max activity detected in mutated hemizygotes and homozygotes was 3.4U/gHb in cord blood, 4.0U/gHb within 24 hours, 2.8U/gHb at day 7, 2.1U/gHb at day 28 and 3.4U/gHb at month 4. All hemizygous and homozygous deficient subjects up to 4 months of life would be identified correctly using the cord-blood threshold (≤4.8IU/gHb) or even the adult threshold (≤4U/gHb) with high sensitivity and specificity. On the other hand, identification of females with heterozygote genotype and deficient or intermediate activity using the cord-blood derived thresholds would have low to very low specificity after 7 days of life.

Neonatal blood goes through extensive changes in the first weeks of life. There are relatively little data describing longitudinal changes from venous cord and neonatal blood in term healthy babies; blood indices are generally similar but tend to be higher in neonatal blood (Greer, Safarulla et al. 2019), particularly for WBC (Scheffer-Mendoza, Espinosa-Padilla et al. 2020) due at least in part to physiological dehydration of infants fed on only colostrum on the first day of life. This is the first study that analysed G6PD activity in cord blood and capillary blood collected up to 7 days of life. Together with the physiologic changes observed in G6PD activity, increased number of WBC and young reticulocytes would have impacted on total detected activity, especially in G6PD deficient and intermediate samples.

A study limitation was the length of follow-up of participants up to 4 months of life. As shown before (Bancone, Poe et al. 2022), G6PD activity does not reach adult levels by 6 months of age and possibly not until one year of age. As a consequence, the Biosensor has not been evaluated yet in infants and children between 4 months and 2 years of age, leaving a diagnostic gap for safe administration of several drugs, including antimalarial radical cure in younger children. While using adult thresholds would probably identify most of the patients with lower activities, correct identification of females with the appropriate residual enzymatic activity to receive safely haemolytic 8-aminoquinolines might require assessment of age specific thresholds.

In conclusion, this study has showed that the “STANDARD G6PD™” test can be used in both cord blood and capillary neonatal blood collected during the first week of life to provide reliable identification of G6PD abnormal neonates who are at increased risk of neonatal hyperbilirubinaemia. Equivalent performance in cord and capillary blood means enhanced use in different contexts and for different clinical needs. Further evaluation outside Thailand (where consistent results have been obtained so far in neonates of Thai, Karen and Burman ethnicity) would further strengthen the generalizability of these findings and support wider use of this useful tool especially in low-resources settings.

## Data Availability

De-identified participant data are available from the Mahidol Oxford Tropical Medicine Data Access Committee upon request from this link: https://www.tropmedres.ac/units/moru-bangkok/bioethics-engagement/datasharing.

## Ethics

The study protocol and its associated documents were approved by the Oxford Tropical Research Ethics Committee (OXTREC 530-23) and the Faculty of Tropical Medicine Ethics Committee (TMEC 23-023), Mahidol University. In addition, the Tak Province Border Community Ethics Advisory Board (T-CAB) was consulted for local feedback and advice.

## Acknowledgements

The authors wish to thank all the mothers for their collaboration and understanding; the study would not have been possible without the hard work and dedication of all SMRU staff involved.

## Supporting information

**Table S1** Mean (SD) haemoglobin levels by day of sampling and instrument

**Table S2** Intraclass coefficient of correlation for G6PD activity between Biosensor and Spectrophotometry

**Table S3** Mean differences haemoglobin (Biosensor – CBC) by sampling day

**Table S4** Mean differences haemoglobin (Biosensor – Hemocue) by sampling day

**Table S5** Mean (SD) values for selected blood indices over time

**TableS6** Summary of Biosensor data in neonates, infants, older children and adults

**Figure S1** Age at sampling of the first capillary blood sample

**Figure S2** Scatter plot of activity by Biosensor in cord blood against capillary sample (<24h of life)

**Figure S3** Scatter plot of activity by Biosensor in cord blood against capillary sample (day 7)

**Figure S4** White blood cells count change over time

**Figure S5** Red blood cells count change over time

**Figure S6** Haemoglobin concentration (by complete blood count) change over time

**Figure S7** Reticulocyte percentage (by complete blood count) change over time

**Figure S8** Bland-Altman plot of G6PD results in cord blood

**Figure S9** Bland-Altman plot of G6PD results in capillary blood collected within 24h of life

**Figure S10** Bland-Altman plot of G6PD results at day7

**Figure S11** Bland-Altman plot of G6PD results at day28

**Figure S12** Bland-Altman plot of G6PD results at M4

**Figure S13** Bland-Altman plot for haemoglobin detected by Biosensor against CBC

**Figure S14** Bland-Altman plot for haemoglobin detected by Biosensor against Hemocue

